# Covid-19 fatality prediction in people with diabetes and prediabetes using a simple score at hospital admission

**DOI:** 10.1101/2020.11.02.20224311

**Authors:** H. Sourij, F. Aziz, A. Bräuer, C. Ciardi, M. Clodi, P. Fasching, M. Karolyi, A. Kautzky-Willer, C. Klammer, O. Malle, A. Oulhaj, E. Pawelka, S. Peric, C. Ress, C. Sourij, L. Stechemesser, H. Stingl, TM. Stulnig, N. Tripolt, M. Wagner, P. Wolf, A. Zitterl, S. Kaser, for the COVID-19 in Diabetes in Austria - study group

**Affiliations:** Clinical Division for Endocrinology and Diabetology, Medical University Graz, Austria; Center for Biomarker Research in Medicine (CBMed), Graz, Austria; Medical Division for Endocrinology, Rheumatology and Acute Geriatrics, Wilhelminen Hospital Vienna, Austria; Clinical Division for Internal Medicine, Endocrinology, Diabetology and Metabolic Diseases St. Vinzenz Hospital Zams, Austria; Clinical Division for Internal Medicine, Konventhospital Barmherzige Brüder Linz, Austria; 4^th^Medical Division with Infectiology, SMZ Süd – KFJ-Hospital Vienna, Austria; Clinical Division for Endocrinology and Diabetology and Metabolic Diseases, AKH Vienna, Austria; Institute of Public Health, College of Medicine and Health Sciences, Al Ain, United Arab Emirates University; 3^rd^ Department and Karl Landsteiner Institute for Metabolic Diseases and Nephrology, Clinic Hietzing, Vienna Health Care Group, Austria; Department for Internal Medicine I, Medical University Innsbruck, Austria; Clinical Division for Cardiology, Medical University Graz, Austria; Department for Internal Medicine I, Paracelsus Medical University, Salzburg, Austria; Clinical Division for Internal Medicine, Hospital Melk, Austria

**Keywords:** prediabetic state, diabetes mellitus, coronavirus infection

## Abstract

**AIM:** We assessed predictors of in-hospital mortality in people with prediabetes and diabetes hospitalized for COVID-19 infection and developed a risk score for identifying those at the highest risk of a fatal outcome.

**MATERIALS AND METHODS:** A combined prospective and retrospective multicenter cohort study was conducted in 10 sites in Austria on 247 people with diabetes or newly diagnosed prediabetes, who were hospitalised for COVID-19. The primary outcome was in-hospital mortality and predictor variables at the time of admission included clinical data, comorbidities of diabetes or laboratory data. Logistic regression analyses were performed to identify significant predictors and develop a risk score for in-hospital mortality.

**RESULTS:** The mean age of people hospitalized (n=238) for COVID-19 was 71.1 ± 12.9 years, 63.6% were males, 75.6% had type 2 diabetes, 4.6% had type 1 diabetes, and 19.8% had prediabetes. The mean duration of hospital stay was 18 ± 16 days, 23.9% required ventilation therapy, and 24.4% died in the hospital. Mortality rate in people with diabetes was numerically higher (26.7%) as compared to those with prediabetes (14.9%) but without statistical significance (p=0.128). A score including age, arterial occlusive disease, CRP, eGFR and AST levels at admission predicted in-hospital mortality with a C-statistics of 0.889 (95%CI: 0.837 – 0.941) and calibration of 1.000 (p=0.909).

**CONCLUSIONS:** The in-hospital mortality for COVID-19 was high in people with diabetes and not significantly different to the risk in people with prediabetes. A risk score using five routinely available patient parameters demonstrated excellent predictive performance for assessing in-hospital mortality.

## INTRODUCTION

After the emergence of the SARS-CoV-2 virus induced COVID-19 ^1^ disease at the end of last year in Wuhan, China, the virus has rapidly spread across the world to achieve pandemic status.

Initial reports from China ^2^,^3^ followed by the US ^4^ and Europe ^5^ showed that the prevalence of diabetes mellitus was as high as 20% of in people being hospitalised for COVID-19. Moreover, the epidemiologic data also suggested that diabetes mellitus is more frequent in people experiencing adverse clinical outcomes ^6^. The prevalence of diabetes was high in people experiencing severe disease and other studies demonstrated higher mortality rates in people with diabetes as compared to non-diabetic cohorts ^7^. In addition, another study also highlighted the high prevalence of prediabetes in people experiencing severe COVID-19 disease. ^8^

Previous research has also shown that people with diabetes face an increased risk of infections that can potentially be explained by impaired phagocytosis by neutrophils, macrophages and monocytes, impaired neutrophil chemotaxis, and bactericidal activity as well as impaired innate cell-mediated immunity ^9^,^10^. Although some observational studies suggest that good glycaemic control might go along with reduced risk for infectious disease, the literature is still conflicting ^9^.

During the COVID-19 lockdown phases in various countries, the question arose, which population groups are particularly at a high risk for severe COVID-19 episodes or death, as those are the ones requiring particular protection and once being affected by the disease, rapid risk stratification in people with disturbed glucose metabolism is critical for further therapy planning or even studies investigating novel treatment approaches. Given the high prevalence of diabetes mellitus in COVID-19, all people with diabetes were initially considered as high-risk population. However, while further studies showed an independent impact of diabetes on the outcome in people with SARS-CoV2 infection, ^7^ also became evident that age and comorbidities, play a major role for the unfavourable outcomes ^11^.

In order to untangle the contribution of diabetes itself and the associated comorbidities, the Austrian Diabetes Association initiated a COVID-19 registry in people with diabetes or prediabetes with the aim to identify those at the highest risk of lethal disease outcome when hospitalized for a SARS-CoV-2 infection. The aim of the current study is to analyse fatality rates in people with diabetes or prediabetes hospitalized for COVID-19 in Austria and to develop an easily applicable score to identify people at highest risk for a fatal outcome within this patient population.

## MATERIALS AND METHODS

### Study Design and Population

We initiated a combined prospective and retrospective, multicentre, and non-interventional cohort study in 10 hospital sites in Austria to collect information on characteristics of people with diabetes and confirmed SARS-CoV-2 infection.

Nationwide ethics approval was obtained from the Ethics Committee of the Medical University of Graz, Austria (EK 32-355 ex 19/20). The study sponsor was the Austrian Diabetes Association.The study started on the 15^th^ of April, for this analysis we used data entered up to the end of June 2020.

Written informed consents were collected wherever possible from living patients. Living patients who were unable to give their consent to participate in this study before discharge, were contacted later for consent to use their clinical data. For patients deceased before consenting, or where consenting to the trial was not feasible, the ethics committee waived the requirement for an informed consent and pseudonymised, retrospective data could be used and included into the analysis as otherwise data would be biased with potentially false low mortality rates.

### Study participants or Inclusion Criteria

People aged 18+ years with confirmed positive throat swab for SARS-CoV-2, confirmed diagnosis of type 1 diabetes, type 2 diabetes, or prediabetes were included in the registry (either known or newly diagnosed). For this analysis we included only those hospitalized for COVID-19. Diabetes mellitus was diagnosed according to the Austrian Diabetes Association ^12^. Prediabetes was defined as an HbA1c between 5.7% and 6.4% (39-46 mmol/mol). ^12^ HbA1c was measured in case increased glucose values were evident in people without known diabetes. No specific exclusion criteria were defined.

### Data collection

Data collection was performed by clinical physicians and study coordinators in 10 participating centres within Austria. Data were collected from their medical files and the clinical laboratory.

This study captured and processed the data using an electronic Case Report Form (HybridForms), which is a validated electronic data capture system with an audit trail and controlled level of access, which was provided by Kapsch BusinessCom and Icomedias, Graz, Austria.

### Study variables

#### Outcome

The primary outcome of this study was in-hospital mortality in patients with diabetes and confirmed diagnosis of COVID-19.

#### Predictors

Demographic information, clinical characteristics, and laboratory findings were collected from the medical record systems of the participating centres. Demographic data consisted of information about age and gender. Clinical characteristics included the classification of diabetes, duration of diabetes, microvascular (diabetic retinopathy and diabetic kidney disease) and macrovascular disease (stroke, myocardial infarction, chronic heart disease, arterial occlusive disease (*i*.*e*. cerebrovascular or peripheral artery disease), other comorbidities of interest (autoimmune disease, cancer, respiratory disease, liver disease, transplantation), and vital signs. Furthermore, current therapy to regulate blood pressure, blood sugar, blood lipids, immunity and pain were recorded. Laboratory data, as far as available from the local lab at the clinical site, included HbA1c, fasting glucose, leucocytes, haemoglobin, estimated glomerular filtration rate (eGFR) (all but one center (MDRD) were using the CKD-EPI formula), high sensitive C-reactive protein (CRP), inflammatory markers, liver function parameters, lipid status, procalcitonin, ferritin, interleukine-6, n-terminal pro brain natriuretic peptide (NT-proBNP), and troponin T. Parameters recorded in the registry are the ones at hospital admission.

### Statistical analyses

All statistical analyses were performed in Stata 16.1 (Texas, US). Qualitative variables were presented as frequency and percentage (%) and quantitative variables as mean ±standard deviation (SD) or median and interquartile range (IQR) as appropriate. Chi-square or Fischer Exact tests were performed to compare qualitative variables and unpaired t-tests or Mann-Whitney U tests to compare normal and non-normal quantitative variables. A p-value of <0.05 was considered statisticaly significant.

### Derivation of risk model

Logistic regression was applied to derive the risk model. The candidate predictors for the model were selected based on their clinical relevance, absence (predictors with <20% of missing data were selected), and p-value ≤0.20 in the univariate logistic regression analysis. The stepwise backward elimination method was applied on candidate predictors to identify predictors for developing the final model. The predictors with p-values ≤0.10 were retained the final model. In addition, interaction effects of various predictors were evaluated in the model.

The risk equation was derived from the final model to predict the log-odds of in-hospital mortality of COVID-19 by summing the product of constant (β_0_) with the product of β coefficients and values of each predictor included in the model. The resulting log-odds were then converted into the probability of in-hospital mortality.

### Performance of risk model

The predictive performance of the risk model was assessed in terms of discrimination and calibration. Discrimination was assessed by calculating the C statistics and calibration was assessed by performing the Hosmer-Lameshaw goodness of fit test and fitting calibration plots of observed versus expected probability of the in-hospital mortality.

### Validation of risk model

The bootstrap method was used for the internal validation of the risk model. The bootstrap samples (n=1000) were drawn from the whole derivation cohort and the model was developed in each bootstrap sample adopting the same methodology of the derivation cohort and using the ‘swboot’ package of Stata. The predictive performance of the risk model was evaluated on bootstrap samples and then on the derivation cohort to get optimism-corrected estimates of the performance of the risk model.

### Development of Nomogram

After generating and validating the risk model, a nomogram was generated from the multivariable logistic regression model using the ‘nomolog’ package of Stata software.

## RESULTS

### Demographics and clinical characteristics

In total, 247 people with diabetes or prediabetes positively tested for SARS-CoV-2 in the participating hospitals in Austria were recorded in the registry, 238 were admitted to the in-patient ward and are part of the current analyses.

The mean age of all participants was 71.1 ± 12.9 years and 63.9% (152) were male. 75.6% (n=180) had established type 2 diabetes, 4.6% (n=11) had type 1 diabetes, and 19.8% (n=47) had prediabetes (table 1). People with prediabetes were older as compared to people with established diabetes mellitus (71.9 ± 12.5 versus 67.6 ± 14.0 years, p=0.043). (table 2). Median HbA1c at admission was 5.9 (IQR 0.3) and median fasting glucose 107 mg/dl (IQR 93) mg/dl in people with prediabetes.

**Table 1.** Comparison and unadjusted odds ratios (95% confidence interval) of characteristics, anthropometric indices, comorbidities, medications, and laboratory parameters with in-hospital mortality in patients hospitalized for COVID-19.

| Characteristics | All |  | In-hospital mortality |  |  |  |
| --- | --- | --- | --- | --- | --- | --- |
|  | N | Statistics | Yes | No | Unadjusted OR (95% CI) | P-Value |
| All, n (%) | 238 | -- | 58 (24.4) | 180 (76.6) | -- | -- |
| **Characteristics** |  |  |  |  |  |  |
| Age – years, Mean ±SD | 238 | 71.1 ±12.9 | 79.8 ±8.8 | 68.3 ±12.8 | 1.66 (1.38 – 1.98)* | <0.001 |
| Sex, n (%) | 238 |  |  |  |  |  |
| Male |  | 152 (63.9) | 36 (62.1) | 116 (64.4) | 1 |  |
| Female |  | 86 (36.1) | 22 (37.9) | 64 (35.6) | 1.10 (0.56 – 2.16) | 0.787 |
| Smoking status, n (%) | 238 |  |  |  |  |  |
| Non-smoker |  | 196 (82.3) | 46 (79.3) | 150 (83.3) | 1 |  |
| Former smoker |  | 38 (16.0) | 12 (20.7) | 26 (14.4) | 1.30 (0.62 – 2.75) | 0.485 |
| Current smoker |  | 4 (1.7) | 0 (0.0) | 4 (2.2) |  |  |
| Body Mass Index – kg/m<sup>2</sup>, Mean ±SD | 114 | 29.1 ±5.7 | 29.2 ±5.8 | 29.0 ±5.7 | 1.01 (0.94 – 1.08) | 0.847 |
| **Vital signs** |  |  |  |  |  |  |
| Systolic BP – mmHg, Mean ±SD | 154 | 132.8 ±21.9 | 134.1 ±25.5 | 132.4 ±20.5 | 1.02 (0.94 – 1.10)<sup>#</sup> | 0.670 |
| Diastolic BP – mmHg, Mean ±SD | 154 | 76.6 ±16.2 | 77.1 ±21.9 | 76.4 ±13.6 | 1.01 (0.91 – 1.03)<sup>#</sup> | 0.790 |
| Pulse – beats/min, Mean ±SD | 137 | 87.3 ±17.4 | 88.2 ±20.9 | 87.0 ±16.1 | 1.02 (0.92 – 1.14)<sup>s</sup> | 0.717 |
| **Diabetes** |  |  |  |  |  |  |
| Type of diabetes, n (%) | 238 |  |  |  |  |  |
| Prediabetes |  | 47 (19.8) | 7 (12.1) | 40 (22.2) | 1 |  |
| Type 1 diabetes |  | 11 (4.6) | 1 (1.7) | 10 (5.6) | 0.46 (0.44 – 4.90) | 0.522 |
| Type 2 diabetes |  | 180 (75.6) | 50 (86.2) | 130 (72.2) | 1.85 (0.66 – 5.16) | 0.240 |
| **Comorbidities** |  |  |  |  |  |  |
| Hypertension, n (%) | 238 | 169 (71.0) | 50 (86.2) | 119 (66.1) | 2.93 (1.25 – 6.90) | **0.014** |
| CHD, n (%) | 238 | 63 (26.5) | 23 (39.7) | 40 (22.2) | 2.14 (1.08 – 4.23) | **0.028** |
| Myocardial infarction, n (%) | 238 | 29 (12.2) | 12 (20.7) | 17 (9.4) | 2.12 (0.87 – 5.13) | 0.097 |
| Heart failure, n (%) | 238 | 30 (12.6) | 15 (25.9) | 15 (8.3) | 3.87 (1.64 – 9.13) | **0.002** |
| Arterial occlusive disease, n (%) | 238 | 38 (16.0) | 18 (31.0) | 20 (11.1) | 4.08 (1.79 – 9.29) | **0.001** |
| Stroke, n (%) | 238 | 20 (8.4) | 8 (13.8) | 12 (6.7) | 2.58 (0.91 – 7.31) | 0.074 |
| Chronic Kidney disease, n (%) | 238 | 55 (23.1) | 24 (41.4) | 31 (17.2) | 3.18 (1.55 – 6.54) | **0.002** |
| Cancer, n (%) | 238 | 37 (15.5) | 12 (20.7) | 25 (13.9) | 1.32 (0.58 – 2.99) | 0.503 |
| Respiratory disease, n (%) | 238 | 48 (20.2) | 15 (25.9) | 33 (18.3) | 1.40 (0.65 – 3.01) | 0.387 |
| Liver disease, n (%) | 238 | 12 (5.0) | 6 (10.3) | 6 (3.3) | 5.11 (1.32 – 19.70) | **0.018** |
| **Medication** |  |  |  |  |  |  |
| Insulin, n (%) | 238 | 52 (21.9) | 12(20.7) | 40 (22.2) | 0.83 (0.37 – 1.82) | 0.636 |
| Other glucose-lowering drugs, n (%) | 238 | 111 (46.6) | 31 (53.5) | 80 (44.4) | 1.39 (0.71 – 2.70) | 0.338 |
| Metformin, n (%) | 238 | 77 (32.3) | 14 (24.1) | 63 (35.0) | 0.55 (0.26 – 1.14) | 0.111 |
| Sulphonylurea, n (%) | 238 | 14 (5.9) | 6 (10.3) | 8 (4.4) | 2.13 (0.63 – 7.15) | 0.220 |
| DPP-4 inhibitors, n (%) | 238 | 42 (17.7) | 15 (25.9) | 27 (15.0) | 1.84 (0.85 – 3.97) | 0.121 |
| SGLT-2 inhibitors, n (%) | 238 | 24 (10.1) | 3 (5.2) | 21 (11.7) | 0.38 (0.10 – 1.46) | 0.158 |
| GLP-1 agonists, n (%) | 238 | 3 (1.3) | 0 (0.0) | 3 (1.7) |  |  |
| Antihypertensive drugs, n (%) | 238 | 169 (71.0) | 46 (79.3) | 125 (69.4) | 1.49 (0.70 – 3.20) | 0.303 |
| ACE inhibitors, n (%) | 238 | 72 (30.3) | 21 (36.2) | 51 (28.3) | 1.47 (0.75 – 2.88) | 0.262 |
| AR blockers, n (%) | 238 | 49 (20.6) | 10 (17.2) | 39 (21.7) | 0.66 (0.29 – 1.50) | 0.319 |
| Beta blockers, n (%) | 238 | 95 (39.9) | 30 (51.7) | 65 (36.1) | 1.81 (0.95 – 3.45) | 0.071 |
| Ca<sup>+</sup> channel blockers, n (%) | 238 | 73 (30.7) | 18 (31.0) | 55 (30.6) | 1.09 (0.55 – 2.15) | 0.811 |
| Central antihypertensive drugs, n (%) | 238 | 10 (4.2) | 3 (5.2) | 7 (3.9) | 1.65 (0.38 – 7.02) | 0.501 |
| Thiazide diuretics, n (%) | 238 | 36 (15.1) | 6 (10.3) | 30 (16.7) | 0.58 (0.23 – 1.46) | 0.247 |
| Loop diuretics, n (%) | 238 | 38 (16.0) | 15 (25.9) | 23 (12.8) | 2.38 (1.14 – 4.95) | **0.020** |
| Mineralocorticoid Receptor Blockers, n (%) | 238 | 20 (8.4) | 7 (12.1) | 13 (7.2) | 4.47 (1.11 – 17.89) | **0.034** |
| Sacubitril, n (%) | 238 | 3 (1.3) | 1 (1.7) | 2 (1.1) | 1.56 (0.14 – 17.54) | 0.718 |
| Glucocorticoid therapy, n (%) | 238 | 10 (4.2) | 5 (8.6) | 5 (2.8) | 3.30 (0.92 – 11.84) | 0.067 |
| Immuno-suppressive therapy, n (%) | 238 | 2 (0.8) | 0 (0.0) | 2 (1.1) | 0.61 (0.03 – 12.89) | 0.751 |
| Oral anticoagulants, n (%) | 238 | 32 (13.5) | 12 (20.7) | 20 (11.1) | 1.64 (0.77 – 3.47) | 0.198 |
| Non-vitamin K oral Anticoagulants, n (%) | 238 | 30 (12.6) | 7 (12.1) | 23 (12.8) | 0.94 (0.38 – 2.31) | 0.888 |
| Ibuprofen therapy, n (%) | 238 | 3 (1.3) | 1 (1.7) | 2 (1.1) | 1.56 (0.14 – 17.54) | 0.718 |
| **Laboratory parameters** |  |  |  |  |  |  |
| Leukocytes – 10<sup>9</sup>/L, Mean ±SD | 236 | 7.2 ±3.2 | 8.1 ±4.3 | 6.9 ±2.7 | 1.07 (0.97 – 1.18) | 0.178 |
| Hemoglobin – mg/dl, Mean ±SD | 238 | 13.3 ±2.2 | 12.9 ±1.9 | 13.4 ±2.3 | 0.90 (0.76 – 1.07) | 0.225 |
| eGFR – mL/min/1.73m<sup>2</sup>, Mean ±SD | 233 | 66.3 ±26.9 | 45.0 ±21.2 | 73.1 ±25.0 | 0.95 (0.94 – 0.97) | **<0.001** |
| Lactate Dehydrogenase – U/L, Mean ±SD | 215 | 331.3 ±188.9 | 330.4 ±169.2 | 331.6 ±194.9 | 0.89 (0.39 – 2.06) | 0.786 |
| AST – U/L, Median (IQR) | 220 | 37.5 (27.5) | 40.0 (31.0) | 37.0 (26.0) | 1.01 (1.00 – 1.02) | **0.029** |
| ALT – U/L, Median (IQR) | 233 | 29.0 (23.0) | 27.0 (23.0) | 31.5 (23.0) | 0.80 (0.48 – 1.34) | 0.396 |
| CRP – mg/dl, Median (IQR) | 232 | 9.9 (14.8) | 16.5 (57.9) | 9.0 (11.6) | 1.35 (1.07 – 1.71) | **0.012** |
| Ferritin – ng/ml, Median (IQR) | 193 | 572.0 (938.0) | 590.0 (897.0) | 568.0 (1019.0) | 0.99 (0.88 – 1.10) | 0.792 |
| Procalcitonin – ng/ml, Median (IQR) | 153 | 0.1 (0.2) | 0.3 (0.5) | 0.1 (0.1) | 1.37 (1.06 – 1.76) | **0.016** |
| IL6 – pg/ml, Median (IQR) | 169 | 50.6 (75.8) | 90.5 (149.4) | 41.8 (57.4) | 2.27 (1.37 – 3.75) | **0.001** |
| Fasting plasma glucose – mg/dl, Median (IQR) | 187 | 127.0 (83.0) | 148.5 (63.0) | 121.0 (88.0) | 0.99 (0.99 – 1.01) | 0.464 |
| HbA1c – %, Median (IQR) | 174 | 6.4 (1.4) | 6.6 (0.8) | 6.4 (1.5) | 0.92 (0.69 – 1.22) | 0.554 |
| HDL Cholesterol – mg/dl, Mean ±SD | 124 | 34.1 ±13.5 | 31.9 ±14.1 | 34.8 ±11.7 | 0.98 (0.94 – 1.02) | 0.312 |
| LDL Cholesterol – mg/dl, Mean ±SD | 123 | 72.4 ±30.8 | 65.1 ±31.6 | 74.8 ±30.3 | 0.98 (0.97 – 1.00) | 0.076 |
| Triglycerides – mg/dl, Median (IQR) | 135 | 116.0 (62.0) | 118.0 (66.0) | 115.0 (63.0) | 1.00 (0.99 – 1.01) | 0.354 |
| NT-proBNP – pg/ml, Median (IQR) | 86 | 542.0 (1408.0) | 1250.0 (2646.0) | 253.0 (471.0) | 1.97 (1.25 – 3.08) | **0.003** |
| TroponinT – pg/ml, Median (IQR) | 126 | 20.0 (32.0) | 42.0 (52.0) | 16.0 (21.0) | 3.63 (1.60 – 8.24) | **0.002** |

OR, Odds Ratio; CI, Confidence Interval; BP, blood pressure; CHD, coronary heart disease; DPP-4, Dipeptidylpeptidase 4; SGLT-2, Sodium dependent glucose transporter-2; GLP-1, Glucagon-like peptide-1; ACE, angiotensin converting enzyme; AR, angiotensin receptor; AST, aspartate aminotransferase; ALT, alanine aminotransferase; IL6, interleukin 6; HbA1c, glycated hemoglobin; HDL, high density lipoprotein; LDL, low density lipoprotein; eGFR, estimated glomerular
filtration rate; NT-proBNP, n-terminal pro brain natriuretic peptide; CRP, C-reactive protein; \* odds for change of 5 years; # odds for change of 5 mmHg; § odds for change of 5 beats/min

**Table 2.**
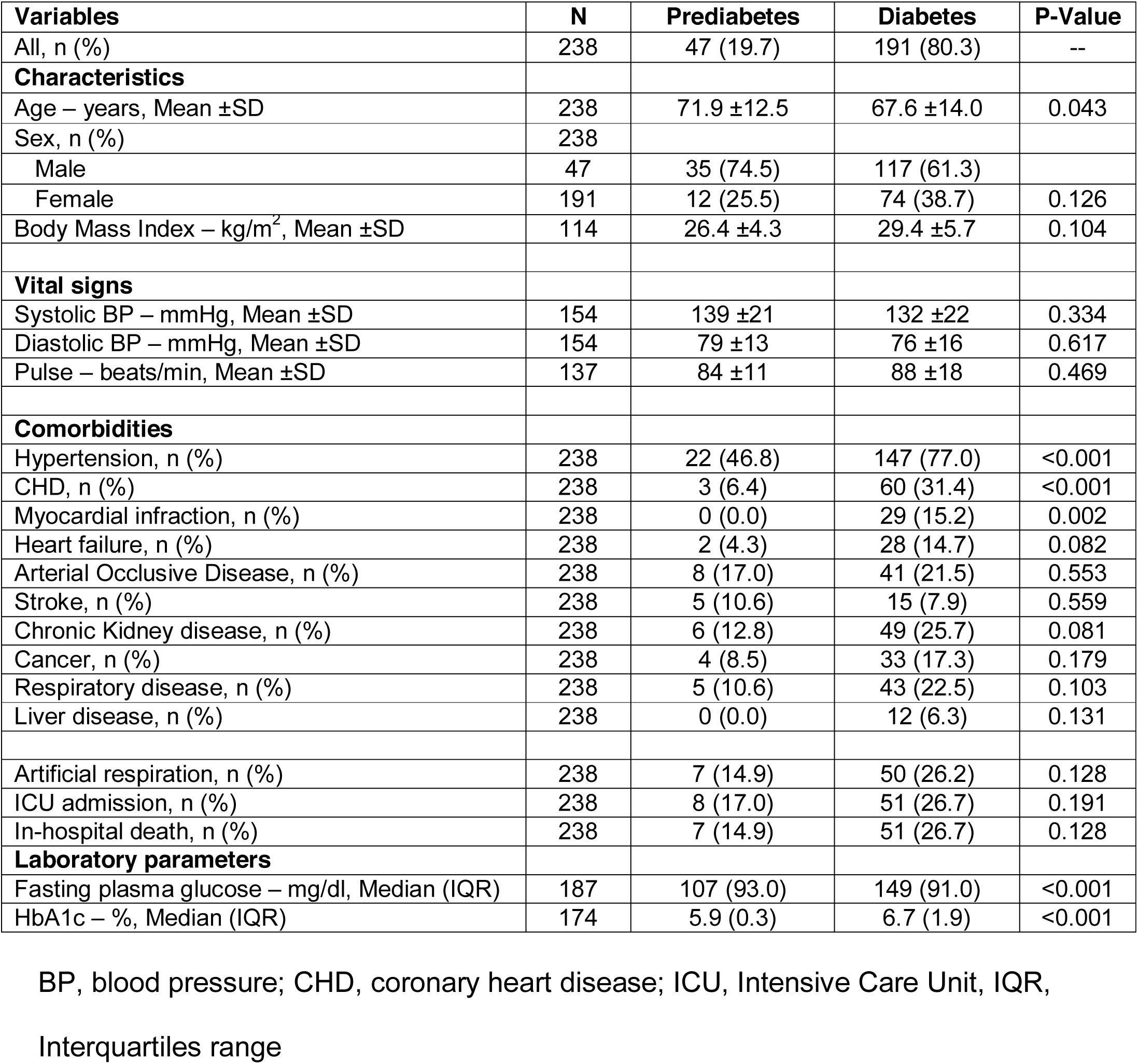
Comparison of characteristics, anthropometric indices, comorbidities, and laboratory parameters by diabetes and prediabetes in patients hospitalized for COVID-19.

| **Variables** | **N** | **Prediabetes** | **Diabetes** | **P-Value** |
| --- | --- | --- | --- | --- |
| All, n (%) | 238 | 47 (19.7) | 191 (80.3) | -- |
| **Characteristics** |  |  |  |  |
| Age – years, Mean ±SD | 238 | 71.9 ±12.5 | 67.6 ±14.0 | 0.043 |
| Sex, n (%) | 238 |  |  |  |
| Male | 47 | 35 (74.5) | 117 (61.3) |  |
| Female | 191 | 12 (25.5) | 74 (38.7) | 0.126 |
| Body Mass Index – kg/m<sup>2</sup>, Mean ±SD | 114 | 26.4 ±4.3 | 29.4 ±5.7 | 0.104 |
| **Vital signs** |  |  |  |  |
| Systolic BP – mmHg, Mean ±SD | 154 | 139 ±21 | 132 ±22 | 0.334 |
| Diastolic BP – mmHg, Mean ±SD | 154 | 79 ±13 | 76 ±16 | 0.617 |
| Pulse – beats/min, Mean ±SD | 137 | 84 ±11 | 88 ±18 | 0.469 |
| **Comorbidities** |  |  |  |  |
| Hypertension, n (%) | 238 | 22 (46.8) | 147 (77.0) | <0.001 |
| CHD, n (%) | 238 | 3 (6.4) | 60 (31.4) | <0.001 |
| Myocardial infraction, n (%) | 238 | 0 (0.0) | 29 (15.2) | 0.002 |
| Heart failure, n (%) | 238 | 2 (4.3) | 28 (14.7) | 0.082 |
| Arterial Occlusive Disease, n (%) | 238 | 8 (17.0) | 41 (21.5) | 0.553 |
| Stroke, n (%) | 238 | 5 (10.6) | 15 (7.9) | 0.559 |
| Chronic Kidney disease, n (%) | 238 | 6 (12.8) | 49 (25.7) | 0.081 |
| Cancer, n (%) | 238 | 4 (8.5) | 33 (17.3) | 0.179 |
| Respiratory disease, n (%) | 238 | 5 (10.6) | 43 (22.5) | 0.103 |
| Liver disease, n (%) | 238 | 0 (0.0) | 12 (6.3) | 0.131 |
| Artificial respiration, n (%) | 238 | 7 (14.9) | 50 (26.2) | 0.128 |
| ICU admission, n (%) | 238 | 8 (17.0) | 51 (26.7) | 0.191 |
| In-hospital death, n (%) | 238 | 7 (14.9) | 51 (26.7) | 0.128 |
| **Laboratory parameters** |  |  |  |  |
| Fasting plasma glucose – mg/dl, Median (IQR) | 187 | 107 (93.0) | 149 (91.0) | <0.001 |
| HbA1c – %, Median (IQR) | 174 | 5.9 (0.3) | 6.7 (1.9) | <0.001 |

BP, blood pressure; CHD, coronary heart disease; ICU, Intensive Care Unit, IQR,
Interquartiles range

**Table 3.** Adjusted coefficients and odds ratios of significant predictors with in-hospital mortality in patients hospitalized for COVID-19.

| Predictor | Adjusted Coefficient<br>(95% CI) | Adjusted OR | P-Value |
| --- | --- | --- | --- |
| Constant | -7.80 | 0.0004 |  |
| Age – years | 0.095 (0.047 – 0.143) | 1.099 (1.048 – 1.153) | <0.001 |
| CRP – mg/dl (log) | 0.012 (0.003 – 0.020) | 1.012 (1.003 – 1.020) | 0.007 |
| eGFR – ml/min/1.73m<sup>2</sup> | -0.036 (-0.055 – -0.0166) | 0.965 (0.947 – 0.983) | <0.001 |
| AST – U/L | 0.020 (0.002 – 0.037) | 1.020 (1.002 – 1.038) | 0.031 |
| Arterial Occlusive Disease |  |  |  |
| No | 1 | 1 | 0.016 |
| Yes | 1.269 (0.234 – 2.305) | 3.558 (1.264 – 10.022) |  |

CI, Confidence Interval; OR, Odds Ratio; AST, Aspartate Aminotransferase; CRP, c-reactive protein; eGFR, estimated Glomerular Filtration Rate

The median duration of hospital stay was 12 (IQR 14) days, whereby almost one-quarter of the cohort was admitted to the Intensive Care Unit (ICU) for the mean stay of 19 ±17 days. Ventilation therapy was needed in 23.9% and in-hospital mortality was 24.4%. Table 1 lists detailed clinical characteristics of all participants and compares those characteristics in people who died in the hospital versus those who were discharged alive. People who died had significantly more often 4 or more known comorbidities at admission (48.3%) as compared to those who survived (12.2%, p<0.001).

With regard to the medication use, no difference between people who survived or died was observed for ACE inhibitors, ARBs, or any glucose lowering medication. Loop diuretics and mineralocorticoid receptor blockers were used more frequently in those who died in the hospital.

### Laboratory findings

The eGFR was significantly lower in people who died in the hospital (45.0 ± 21.2 versus 73.1 ± 25.0, p<0.001). Moreover, CRP levels were significantly higher in people being admitted to the ICU and in those who died, as were procalcitonin, interleukin-6 levels, or NT-proBNP levels.

### Outcomes analyses and risk model

Table 2 displays the adjusted odds ratios of significant predictors of in-hospital mortality identified within our cohort. Besides the age, the presence of arterial occlusive disease, CRP levels, eGFR, and AST levels were significant predictors of in-hospital mortality. No significant differences in the odds of in-hospital mortality were observed with respect to prediabetes and type 1 and type 2 diabetes.

Derived from the logistic regression model and the above-identified parameters, we developed a risk score for in-hospital mortality. The nomogram in figure 1 displays the included parameters and assigns a score value to each of them, either in a categorical or continuous manner. In the derivation cohort, the model achieved the area under the curve (AUC) or C-statistic of 0.889 (95%CI: 0.837 – 0.941) and calibration of 1.000 (Hosmer-Lameshow test p = 0.909). In internal validation using the bootstrapping method, the C-statistic was 0.893 (95%CI: 0.801 – 0.959) and calibration was 0.930 (Hosmer-Lameshow test p = 0.918) (supplementary figure 1).

**Figure 1.**
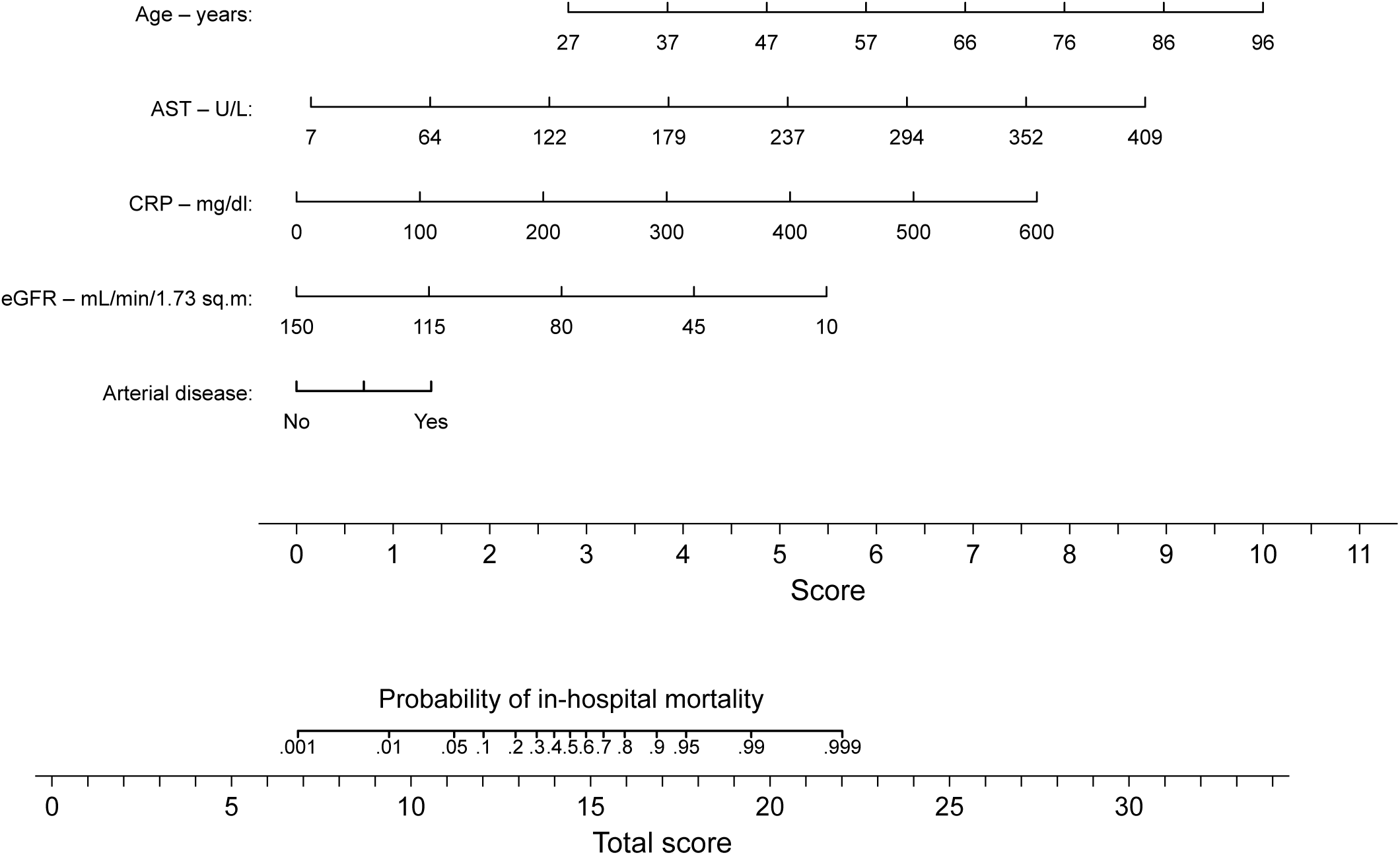
Nomogram for predicting in-hospital mortality in patients hospitalized for COVID-19. AST, Aspartate Aminotransferase; CRP, c-reactive protein; eGFR, estimated Glomerular Filtration Rate Risk is given as probability and to improve readability we have omitted the second decimal place (e.g. 0.90 is written as 0.9). In order to estimate the fatality risk for a patient with diabetes or prediabetes at hospital admission, one need to find the corresponding score points for each of the five clinical characteristics and sum them up. The scale at the bottom gives the probability of in-hospital mortality corresponding to the calculated score.

## DISCUSSION

In our cohort of people with established diabetes and prediabetes, hospitalized for COVID-19 in Austria, the in-hospital mortality was as high as 24.4%. We did not observe a statistically significant difference in people with type 1 and type 2 for mortality, although the number of people with type 1 diabetes was only eleven. More interestingly, the mortality rate in those having prediabetes was numerically lower (14.9%), albeit not statistically significant in comparison to people with type 2 diabetes.

With the identified predictors for in-hospital mortality, being age, presence of arterial occlusive disease, AST, eGFR, and CRP levels at admission we developed a simple clinical score to identify those people at highest risk of fatal outcome.

Earlier this year data on the high prevalence of diabetes in people hospitalized for COVID-19 and in particular severe episodes of the disease emerged throughout the world ^2,3,4,5^. Later, more thorough analyses adjusting the results for co-variates still identified diabetes as a significant risk factor for fatal outcomes ^7^.

The CORONADO study was the first larger dataset investigating people with diabetes hospitalized for COVID-19 in France. ^11^ Similar to our study, they showed that HbA1c at admission was not a significant predictor of outcome in this patient cohort, while a UK analysis suggested a higher mortality rate in people with higher HbA1c levels, both in type 1 and type 2 diabetes. ^13^ A recent Italian dataset demonstrated glucose at admission as a predictor for disease severity and prognosis, however admission glucose seems to mainly reflect the inflammatory response rather than the quality of the pre-COVID-19 glycemic control ^14^. While other publications deal with recommendations for glucose lowering at home, in the hospital setting or around surgery, our data do not look into this important aspect. ^15,16^

In contrast to our data, BMI was a predictor of mortality in the British and the French study; hence this might be a matter of sample size in our study. In addition, the French cohort identified age, obstructive sleep apnoea syndrome, or micro- and macrovascular complications as predictors of adverse outcomes. In terms of laboratory parameters, similar to our findings, AST and CRP are directly and eGFR inversely related to mortality. In addition, a recent Chinese dataset suggested CRP as one major predictor of mortality in people with diabetes ^17^. In line with the CORONADO data, we also did not find a difference in mortality between people with type 1 and type 2 diabetes being hospitalized. In a UK dataset from the NHS, the adjusted odds ratios (for age, sex, deprivation, ethnicity, and geographical region) for in-hospital related COVID-death were higher in people with type 1 diabetes than type 2 diabetes ^18^. However, these data are difficult to interpret, as the analyses were not adjusted for additional confounding factors like diabetes duration and the presence of comorbidities.

Recently Klein et al. analyzed data from an intensive care unit in Austria, where they identified in 36.3% a previously unknown prediabetes ^8^. Also in our cohort, 47 people (19.7%) had prediabetes diagnosed according to the admission HbA1c and their outcome was not different as compared to those with manifest diabetes mellitus, suggesting that also this prediabetic state has an inverse impact on COVID-19 disease progress.

As countries have different approaches to tackle the SARS-CoV-2 pandemic with various hospitalization strategies, outcome figures vary considerably between the countries. While the in-hospital mortality rate in the CORONADO study was 10.6% (9), the mortality rate in people with diabetes hospitalised for COVID-19 was almost a quarter in our study. Hence, we believe it is important to further study country-based mortality rates and to put them into context to the national COVID-19 strategy.

Also in our database, no specific glucose lowering drug was associated with increased or reduced risk for in-hospital death.

For a clinician, simple and easily applicable risk stratification for people admitted to the emergency room is helpful for triaging reasons and the planning of further care. Moreover, this risk stratification is also an important tool to design clinical trials for therapeutic agents, as it is likely that they will have different effects in different risk groups. Therefore, we propose a simple risk score based on age, the presence of arterial occlusive disease, the CRP, AST, and eGFR levels. With an AUC of more than 0.8, this score looks promising, but we were only able to validate it internally by bootstrapping technique and it clearly needs external validations before a clinical application could be considered.

One limitation of our study is the sample size of 238 subjects. However, given the total population of Austria of less than 9 million people and the importance of having in-hospital mortality data in people with diabetes available for as many as possible countries, these data are of importance. Another limitation is the lack of comparison data in people without diabetes from Austria, hospitalized for COVID-19. In addition, due to the pragmatic design we do not have a ful dataset on all laboratory parameters of interest available in this pragmatic registry. Hence, we decided to use only those laboratory parameters in the risk score model, which were available in more than 80% of participants. Sensitivity analyses including further laboratory parameters (even if the frequency was less than 80%) did not change the predictive performance of the score substantially. Given that HbA1c is not routinely measured in all people admitted to the hospital, prediabetes was likely be underdiagnosed in the overall cohort of people having COVID-19, which needs further investigation.

While a strength of this publication is the data on people with prediabetes and COVID-19 and the idea of summarizing the risk parameters into a simple clinical score, the limitation is the lack of external validation of this score, which is of importance for its potential usage in routine care.

Our data show high in-hospital mortality in people with diabetes and prediabetes in Austria. A simple 5 parameter risk score could help in identifying people at the highest risk of fatal outcome but needs further validation in other cohorts.

## Data Availability

The Austrian Diabetes Association has full access to the dataset and access can be granted upon request.

## Acknowledgements

We would like to thank Kapsch Austria, Icomedias and Microsoft Austria for the programming of the electronic case report form and the provision of secure data storage space. We thank Andrew Spencer for language editing.

## Duality of interest

HSourij received unrestricted research grants from AstraZeneca, Boehringer Ingelheim, Eli Lilly, MSD, NovoNordisk and Sanofi. HSourij received speaker’s honoraria from Amgen, AstraZeneca, BMS, Boehringer Ingelheim, Eli Lilly, MSD, NovoNordisk and Sanofi.

SK received unrestricted research grants from Boehringer Ingelheim and MSD (CD Laboratory for Metabolic Crosstalk). SKaser received speaker’s honoraria from AstraZeneca, Boehringer Ingelheim, Eli Lilly, MSD, NovoNordisk and Sanofi. CC received speaker’s honoraria from AstraZeneca, Boehringer Ingelheim, Eli Lilly, MSD, NovoNordisk and Sanofi.

HStingl received an unresctricted research gant from Boehringer Ingelheim. HStingl received speaker’s honoraria from Amgen, AstraZeneca, Boehringer Ingelheim, Eli Lilly, Merck Sharp & Dohme, NovoNordisk, Novartis, and Sanofi Aventis and Servier. CR received speaker’s honoraria and congress support from AstraZeneca, NovoNordisk and Sanofi.

All other authors declare to have no conflichts of interest with regard to this manuscript.

## Author Contributions

HSourij and SK conceived the study, HSourij, NT, CS wrote the protocol and designed the eCRF. HSourij, BA, CC, MC, PF, MK, AKW, CK, OM, EP, SP, CR, CS, LS, HStingl, TS, NT, PW, AZ, KS collected the data. FA and AO performed the statistical analyses. HSourij, CS, NT and FA wrote the first draft of the manuscript and all authors revised the manuscript. HSourij and FA are the guarantors of the data.

**COVID-19 in diabetes in Austria study group**

Medical University Graz, Austria:

*Harald Sourij, Norbert J. Tripolt, Caren Sourij, Farah Abbas, Oliver Malle, Julia Mader*

Medical Division for Endocrinology, Rheumatology and Acute Geriatrics, Wilhelminen Hospital Vienna, Austria:

*Peter Fasching, Gersina Rega-Kaun, Kadriye Aydinkov-Tuzcu, Alexander Bräuer, Brigitte Bernhardt*

Clinical Division for Internal Medicine, Endocrinology, Diabetology and Metabolic Diseases St. Vinzenz Hospital Zams, Austria:

*Christian Ciardi, Marc Schaber, Anna Schapfl, David Fiegl*

Clinical Division for Internal Medicine, Konventhospital Barmherzige Brüder Linz, Austria:

*Martin Clodi, Carmen Klammer, Michael Resl, Matthias Heinzl, Roland Feldbauer, Johannes Pohlhammer*

4^th^Medical Division with Infectiology, SMZ Süd – KFJ-Hospital Vienna, Austria Mario Karolyi, Erich Pawelka

Clinical Division for Endocrinology and Diabetology and Metabolic Diseases, AKH Vienna, Austria:

*Alexandra Kautzky-Willer, Peter Wolf*

Clinical Division for Cardiology, Medical University Graz, Austria:

Department for Internal Medicine I, Paracelsus Medical University, Salzburg, Austria:

*Lars Stechemesser, Michael Schranz*

Clinical Division for Internal Medicine, Hospital Melk, Austria:

*Harald Stingl, Michael Wagner, Reinhard Würfel*

3^rd^ Medical Department and Karl Landsteiner Institute for Metabolic Diseases and Nephrology, Clinic Hietzing, Vienna Health Care Group, Austria

*Thomas M. Stulnig, Slobodan Peric, Andreas Zitterl*

Department for Internal Medicine I, Medical University Innsbruck, Austria:

*Susanne Kaser, Claudia Ress*

## Prior Presentations

**none**

## Preprent publication

This study will be published on https://www.medrxiv.org as a preprint.

