## Supplemental Figure 1 for "Covid-19 fatality prediction in people with diabetes and prediabetes using a simple score at hospital admission"

**Supplemental Material**

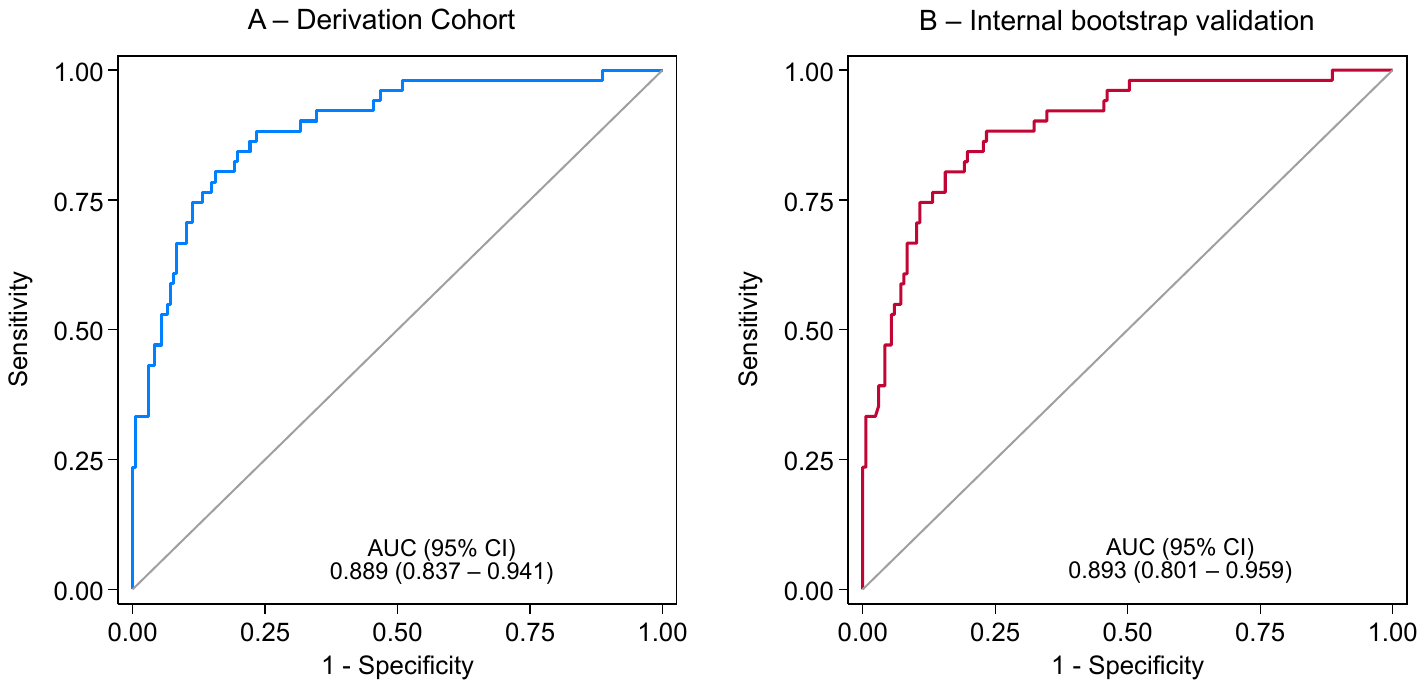

**Supplemental Figure 1 –** ROC curves: A – Derivation cohort, B – Internal bootstrap validation sample
