## Supplemental Figure 2 for "Covid-19 fatality prediction in people with diabetes and prediabetes using a simple score at hospital admission"

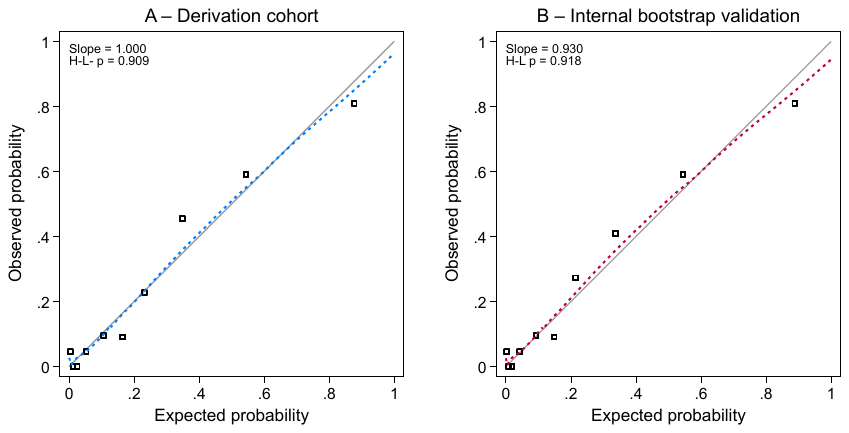

**Supplemental Figure 2 –** Calibration plots: A – Derivation cohort, B – Internal bootstrap validation sample H-L p: Hosmer-Lameshow test p-value
